# Stroke knowledge, perceptions and practices: A large-scale questionnaire in high-risk patients in Bogotá, Colombia (2023 to 2024)

**DOI:** 10.1101/2025.10.07.25337530

**Authors:** Luisa F. Alviz, Juan D. Oyola, Javier Perez, Mariana Medina, Jaime Rodriguez, Juand D. Martinez-Lemus, John Benavides, Carlos Martinez, Claudio Jimenez

**Author notes:** Correspondence to: Claudio Alejandro Jimenez Monsalve, MD Neurology department, Hospital Simón Bolivar Subred Integrada de Servicios Norte, Bogotá, Colombia.

## Abstract

**Objective:** To identify and describe the factors related to the knowledge, perceptions, and practices regarding emergency care for stroke among patients with risk factors such as diabetes and hypertension.

**Background:** Health literacy in stroke implies the ability of non-medical individuals to recognize stroke symptoms and make timely decisions, which is a key factor influencing outcomes in patients with this condition. No previous study has evaluated the knowledge and perceptions of the general population about stroke in Bogotá.

**Methods:** A cross-sectional study was conducted between October 2023 and April 2024. A validated questionnaire surveyed 5402 individuals in Bogotá.

**Results:** Lower stroke symptom knowledge was associated with male sex (OR = 1.26, 95% CI (1.12, 1.42); p < 0.001), low socio-economic strata (OR = 2.20, 95% CI (1.85, 2.60); p < 0.001), and lower educational level (OR = 1.59, 95% CI (1.38, 1.82); p < 0.001). Furthermore, a low educational level was correlated with a higher risk of choosing inadequate courses of action in the event of having an acute stroke. This included taking home remedies (OR = 1.52, 95% CI (1.30, 1.79); p < 0.001) or going to the pharmacy (OR = 1.66, 95% CI (1.36, 1.93); p < 0.001).

**Conclusions:** Factors such as poor recognition of stroke symptoms, failure to identify it as an emergency, and socioeconomic disparities contribute to delays in seeking timely treatment. Addressing knowledge gaps and socioeconomic disparities is crucial for improving outcomes in stroke patients.

## Introduction

According to the World Health Organization (WHO), approximately 15 million individuals experience an Ischemic Stroke (IS) each year. This results in 6 million deaths and leaves 5 million with disabilities. (1) 160.4 million disability-adjusted life years (DALYs) are attributed to IS, ranking this condition as one of the primary causes of mortality and disability worldwide. (2) These statistics are particularly concerning given that stroke is a condition that can be effectively treated during the first hours of symptom onset. Furthermore, it is largely preventable through population-level interventions targeting modifiable risk factors. According to the INTERSTROKE study, up to 90.7% of IS cases could be prevented by addressing 10 modifiable risk factors. The most significant of these included hypertension (HTN), dyslipidemia, obesity, psychosocial stress, smoking, alcohol consumption, and type 2 diabetes mellitus (DM). (3)

In Colombia, there is an absolute increase in the number of stroke cases, driven by population growth, increased life expectancy, and improved detection of stroke in medical services. (4) It remains unclear whether the reduction in mortality has had any impact on the overall number of patients living with stroke-related disabilities.

Another critical factor impacting stroke outcomes is health literacy, which encompasses understanding the disease, recognizing its symptoms, and promptly seeking medical care, among others. (5) A study conducted in 2019 at third-level hospitals in Dhulikhel, Nepal, highlighted the significant gaps in stroke awareness. The study reported that 60% of participants did not know stroke risk factors, only 16% identified “sudden paralysis on one side of the body” as a warning sign, and 64% were unaware of available stroke treatments (n = 273). (6) Similarly, a 2022 study conducted in Al-Ahsa, Saudi Arabia, found that more than half of the participants (56.9%) lacked knowledge of stroke risk factors, warning signs, and prevention strategies (n = 202). However, older adults (aged 50–65) with higher socioeconomic and educational backgrounds, as well as those with a family history of stroke, demonstrated greater awareness, suggesting that the level of health literacy is related to social and economic factors. (7) Moreover, a study conducted in Manizales, Colombia, interviewed 213 individuals, finding that only 11.7% of participants stated they would go to the emergency room if they experienced symptoms of an acute ischemic stroke. (8) Currently, no studies have examined the knowledge, perspectives, and prehospital decision-making regarding stroke presentation in Bogotá.

Therefore, the primary objective of this study is to describe the factors associated with knowledge, perceptions, and practices regarding emergency care for stroke in patients diagnosed with type 2 DM or systemic arterial HTN, treated at hospitals across Bogotá during 2023–2024. The findings of this study will contribute to building evidence that reinforces the need for preventive public health strategies against stroke.

### Methodology

#### Definitions and concepts

In Colombia, the socioeconomic classification system categorizes the population based on income levels, living conditions, and the physical characteristics of homes and their surroundings. (9) The six-level stratum system influences healthcare costs and subsidies for transport and housing, with Stratum 1 denoting the lowest income and living conditions, and Stratum 6 the highest. The system is designed to promote equity by ensuring that higher strata contribute more to service costs, while lower strata benefit from subsidies. (10)

Another key aspect to consider is the Colombian education system. It is divided into two main levels: basic and higher education. Basic education begins with elementary school at age six and lasts five years. Students then progress to secondary school, which includes six years of study, often referred to as high school. Higher education includes universities and technical institutes, offering various degrees based on career goals. The highest academic achievement is the doctoral degree. (11)

In Bogotá, the organization of medical centers follows the structure of Health Lending Institutions (Instituciones Prestadoras de Salud, IPS). These institutions operate under the public health network or private entities. The public health network in Bogotá is divided into four subnetworks: north, south, southwest, and center-east [**Fig 1**]. As of 2016, the city recognized 22 hospitals and 142 health centers. Between 2020 and 2024, the administration added seven more hospitals and 20 health centers. (11,12) However, information on how many facilities are equipped to perform pharmacological thrombolysis or mechanical thrombectomy for ischemic stroke (IS) remains unavailable.

**Figure 1.**
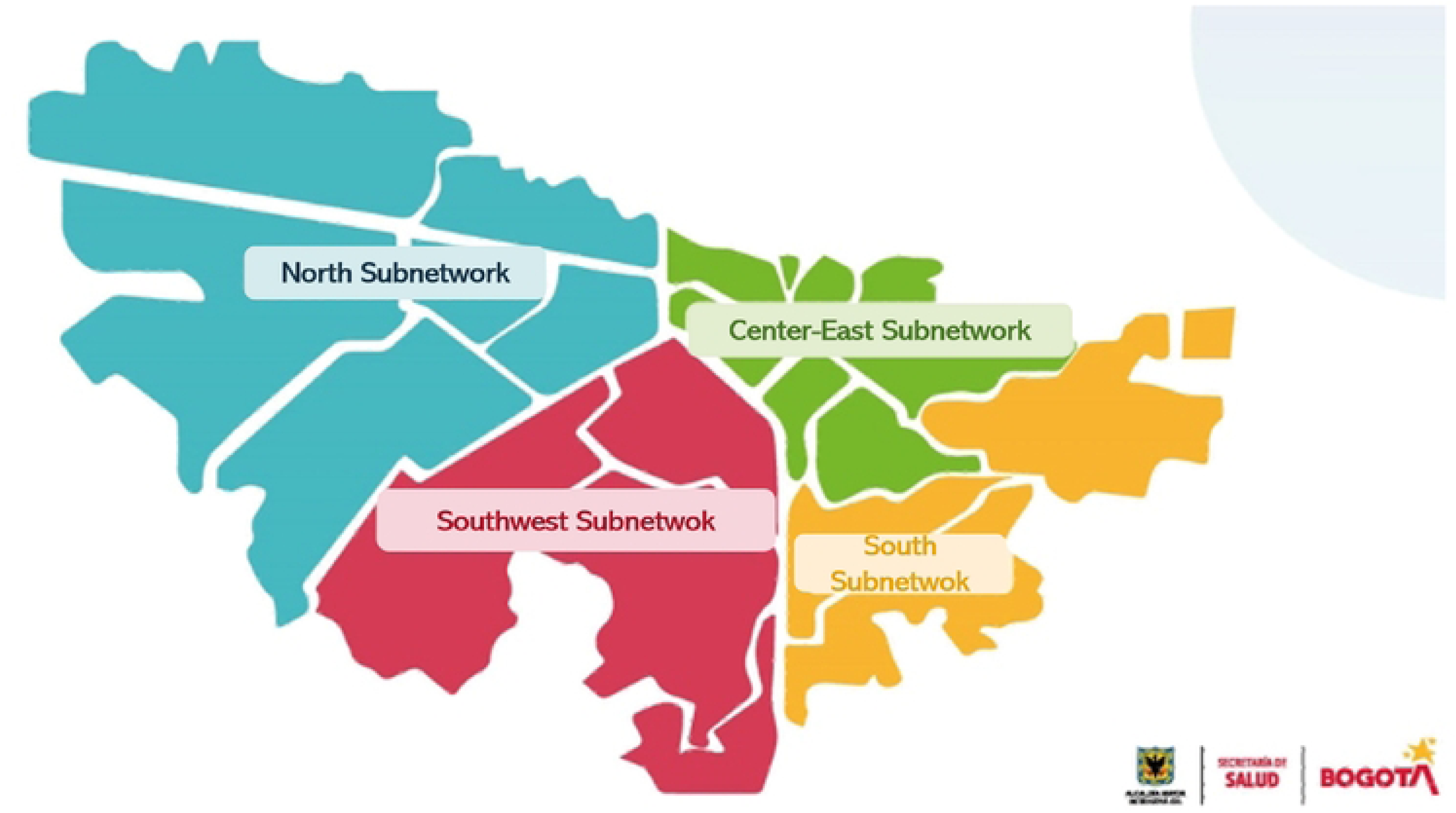
Distribution of the four Integrated Health Subnetworks in Bogotá. This figure shows a representative map of Bogotá City divided by the distribution of health services, which are organized by subnetworks.

#### Study Design, Population, and Eligibility Criteria

An analytical cross-sectional study was conducted among patients living in Bogotá, Colombia, from October 2023 to April 2024. Patients who met the established criteria and attended University Hospital Fundación Santa Fe de Bogotá (UHFSFB) or any hospital within the governmental health subnetworks of the North, South, and Center-East [Fig 1] were included.

#### Inclusion criteria

Patients older than 18 years diagnosed with type 2 DM or HTN who have been living in Bogotá, Colombia, for at least one year.

#### Exclusion criteria

Patients with any prior history of stroke, cerebrovascular event, or any disease that compromises the mental sphere, such as dementia or any degree of intellectual disability.

#### Sample size determination

A stratified non-probability sampling approach by community areas was used. The estimated sample size was calculated using population data obtained from projections by SaluData, with an expected proportion of 24% for HTN (13), which resulted in 1,442,929 individuals. The sample size was 5,402, with a confidence level of 95% and a standard error (precision) of 1.18%.

#### Data collection Tool

To develop the data collection tool, we followed a multi-step methodology. First, a systematic review was conducted following the Preferred Reporting Items for Systematic Reviews and Meta-Analyses (PRISMA) guidelines (14), to address the formulated PICO (Population, Intervention, Comparator, and Outcome) question, detailed in **Supplemental Material S1**. The literature search was constructed using Medical Subject Headings (MeSH) and Descritores em Ciências da Saúde (DeCS) terms, ensuring comprehensive retrieval of relevant studies. Key concepts related to chronic diseases, stroke, and health knowledge, attitudes, and practices were combined using Boolean operators. Literature was captured, and after qualitative and quantitative analysis, 17 articles were deemed eligible for extraction. These articles provided the foundation for constructing the data collection tool. The tool was internally validated by a panel of neurologists and epidemiologists, who extensively evaluated its content. Subsequently, seven pilot tests were conducted with master’s students in Epidemiology to identify and implement potential improvements.

Cronbach’s Alpha Coefficient was calculated to evaluate the internal reliability of the data collection tool, yielding a value of 0.9 for knowledge related to acute stroke and 0.3 for practices concerning stroke.

#### Data collection

Data collection was conducted from October 1, 2023, to April 30, 2024 through phone calls or in person at diverse hospitals from Bogotá. On-site data was collected from patients who met the criteria and attended any of the participating hospitals (including outpatient and emergency department waiting areas, inpatient service, dialysis units, etc).

Information from patients registered in the non-communicable chronic diseases databases from UHFSFB and the North, South, and Center-East health subnetworks was obtained. These databases were filtered to obtain the most up-to-date information on patients with an established diagnosis of type 2 DM or HTN who attended follow-up appointments in general medicine, internal medicine, or primary care. Phone numbers for these patients were identified, and each patient was contacted to administer the questionnaire. After explaining the study’s purpose, potential indirect benefits, ensuring confidentiality, and the right to decline participation, informed consent was obtained. A questionnaire was then administered to those who agreed to participate. Responses were collected via Google Forms, which facilitated the compilation and management of the data.

Weekly meetings were held to evaluate the preliminary data collection and monitor the quality and quantity of questionnaires administered each week. Additionally, random, periodic control samples were selected to verify the accuracy of the collected data. This process involved contacting patients already registered in our database to confirm that the questionnaire had been administered correctly. It helped identify any inconsistencies, allowing for the removal of erroneous data and ensuring high-quality standards in the collected information.

#### Data Analysis

Ordinal and nominal variables were described through proportions. Continuous variables, after performing Kolmogorov-Smirnov normality tests, were described with medians and the interquartile range since there was no normal distribution in any of the variables in the population. To compare sociodemographic factors, a social Strata of 2 or lower was defined as a low socioeconomic strata, an education level of elementary school or less was defined as a low educational level, and an income of 1 million Colombian pesos (Colombian legal tender) per month or less was defined as low income. It is noteworthy that some wealthy individuals reside in lower socioeconomic strata neighborhoods, especially in rural areas, which is a failure in the implementation of the strata system. This is a difficulty when addressing social inequalities and aiding resource allocation in the country. (15,16) This justifies the necessity of conducting a separate analysis of knowledge and practices regarding stroke by income and socioeconomic strata.

Then, a bivariate analysis was performed, comparing the variables with the place of occurrence using a chi-square test. Statistical significance was defined as a p-value < 0.05. Statistical analysis was performed with SPSS software, version 25.

The association between the factor and knowledge was estimated using binomial logistic regression with the exponent of beta as a measure of association and a 95% confidence interval. All models were adjusted for sex, age, and diagnosis of HTN. Both adjusted and unadjusted models are presented in the tables. The goodness of fit for each adjusted model was calculated using the Hosmer-Lemeshow test.

#### Ethical Consideration

Ethical approval was obtained from the ethical review committee of each of the participant institutions: UHFSFB and hospitals within the governmental health subnetworks of the North, South, and Center-East. Written informed consent was obtained from each participant, and confidentiality was ensured throughout the data collection, processing, and analysis.

## Results

A total of 6,050 patients with HTN and/or type 2 DM completed the questionnaire, of whom 5,402 met the study’s inclusion criteria. Of those, 5381 had complete information registered in the database available for the analysis. The median age of participants was 67 years (IQR: 59.68 - 74.94), with 46.3% being female and 53.7% male. Median Body Mass Index (BMI) was 26.9 (IQR, 24.65-29.30). Data were collected from individuals receiving care across the north, south, and center-east health subnetworks. The majority of participants (81.6%) were from low socioeconomic strata and enrolled in the subsidized health insurance regimen (92.4%). Notably, 55.3% of the participants had completed only elementary school.

Included individuals were primarily diagnosed with HTN, accounting for 92% of the cases, with no significant differences in sex distribution (p=0.154). Type 2 DM was reported in 32.1% of cases, with a higher prevalence among men (34%) compared to women (p=0.007). Cholesterol or triglyceride abnormalities were reported by 39.4% of participants, with greater awareness among women compared to men (44.5% vs. 33.5%, p<0.001). Lastly, smoking was more common among men, with a prevalence of 5.9% compared to 2.2% in women (p<0.001) [**Table 1**].

**Table 1.** Questions about self-reported comorbidities.

| **Table 1. Questions about self-reported comorbidities** |  |  |  |  |
| --- | --- | --- | --- | --- |
| **Comorbidities** | **Women**<br>**n=2889 (%)** | **Men**<br>**n= 2492 (%)** | **Total**<br>**n = 5381 (%)** | **p-value\*** |
| **Hypertension**<br>**n = 5381** | 2678 (92.7) | 2284 (91.7) | 4962 (92.2) | 0.154 |
| **Diabetes**<br>**n = 5381** | 882 (30.5) | 847 (34) | 1729 (32.1) | 0.007 |
| **Dyslipidemia**<br>**n = 5379** | 1285 (44.5) | 834 (33.5) | 2119 (39.4) | < 0.001 |
| **Cigarette smoking**<br>**n = 5381** | 64 (2.2) | 146 (5.9) | 210 (3.9) | < 0.001 |
| **\*Corresponding statistic test, significant at p <0.05.** |  |  |  |  |

Among those surveyed, 89.6% recognized stroke as a disease that can lead to disability, and 91.1% associated it with the risk of death. A total of 32.5% of participants reported being unaware of any symptoms related to IS, with this lack of knowledge being more prevalent among men (36.5%) compared to women (29.1%, p<0.001). In terms of practices and decision-making in response to a potential stroke, 26.6% of participants disagreed with activating the emergency medical system by calling the emergency hotline—an action recommended by international guidelines and the Colombian health system. Notably, women were more likely to support calling the emergency hotline than men (p=0.004). When it came to seeking medical care, most patients preferred scheduling an outpatient appointment via telephone, with 52% stating they would contact their insurance provider first. This trend was significantly higher among men compared to women (p<0.001) [**Table 2**].

**Table 2.** Practices regarding emergency care for stroke.

| **Table 2. Practices regarding emergency care for stroke** |  |  |  |  |
| --- | --- | --- | --- | --- |
| **Practice** | **Women**<br>**n=2889 (%)** | **Men**<br>**n= 2492 (%)** | **Total**<br>**n= 5381 (%)** | **p-value\*** |
| **Call emergency number 123** | 2073 (71.8) | 1875 (75.2) | 3948 (73.4) | 0.004 |
| **Make a medical appointment** | 1714 (59.3) | 1085 (43.5) | 2799 (52) | < 0.001 |
| **Go to the pharmacy** | 507 (17.5) | 617 (24.8) | 1124 (20.9) | < 0.001 |
| **Take a home remedy** | 514 (17.8) | 485 (19.5) | 999 (18.6) | 0.116 |
| **\*Corresponding statistic test, significant at $p < 0.05$** |  |  |  |  |

Regarding the odds of knowledge about the symptoms of IS, a statistically significant association was found between limited knowledge of IS symptoms and being male, having low educational attainment, and residing in lower socioeconomic conditions [Fig. 2].

**Figure 2.**
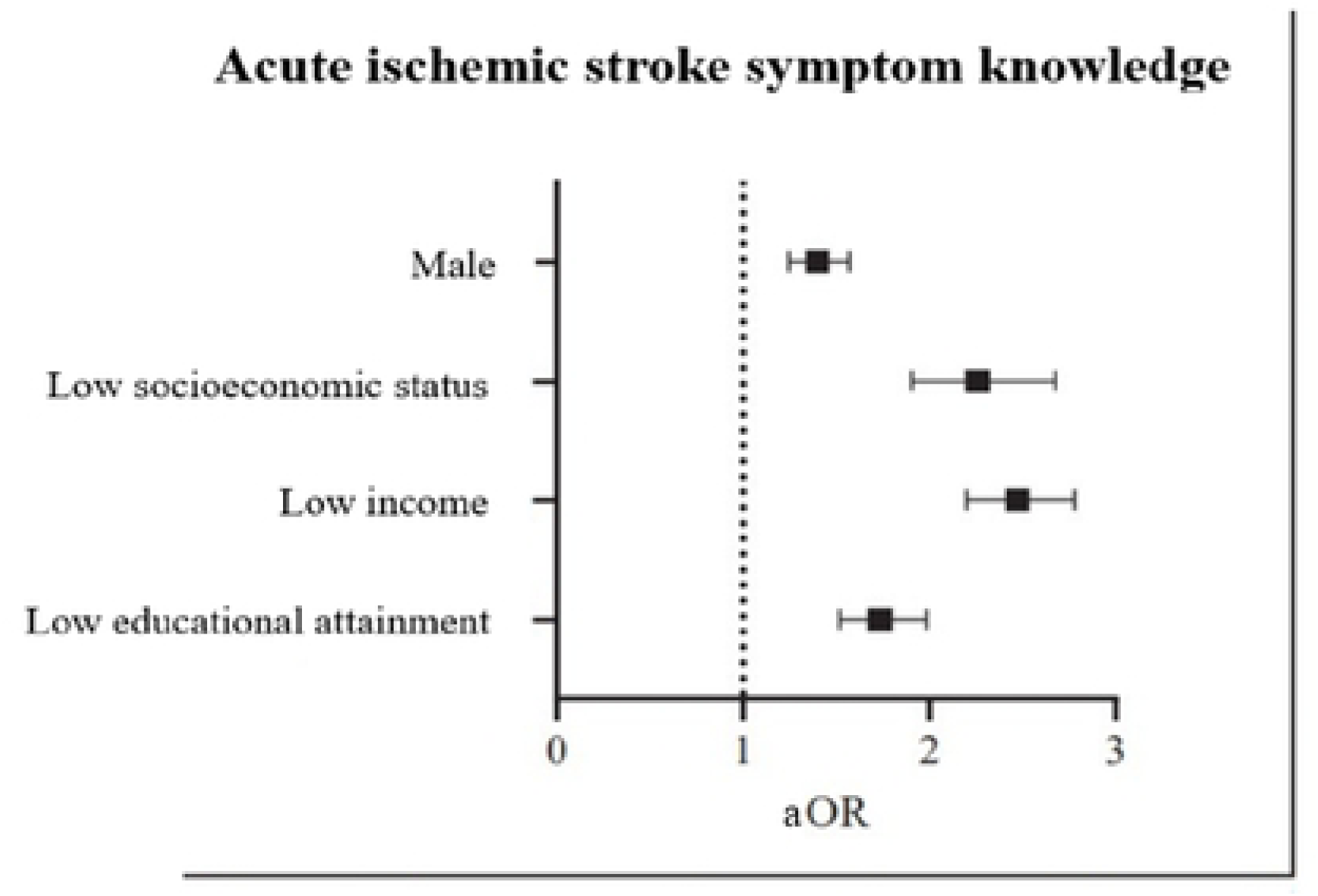
Predictive Factors of Stroke Symptom Knowledge. Summary of estimation of the association between acute ischemic stroke symptom knowledge among participants and sociodemographic variables. Odds Ratio (aOR) was adjusted by variables of age and past medical history of systemic arterial hypertension.

Associations were observed between opting for emergency care practices, such as using home remedies or visiting the pharmacy instead of seeking urgent care at a health center, and being male or living in lower socio-demographic conditions [Fig. 3]. In contrast, the odds of making a medical appointment were lower among men, individuals with lower socioeconomic status, those with lower incomes, and those with lower educational attainment. Similarly, the odds of calling emergency numbers were lower among men, individuals with low educational attainment, and those with lower incomes [Fig. 3].

**Fig 3.**
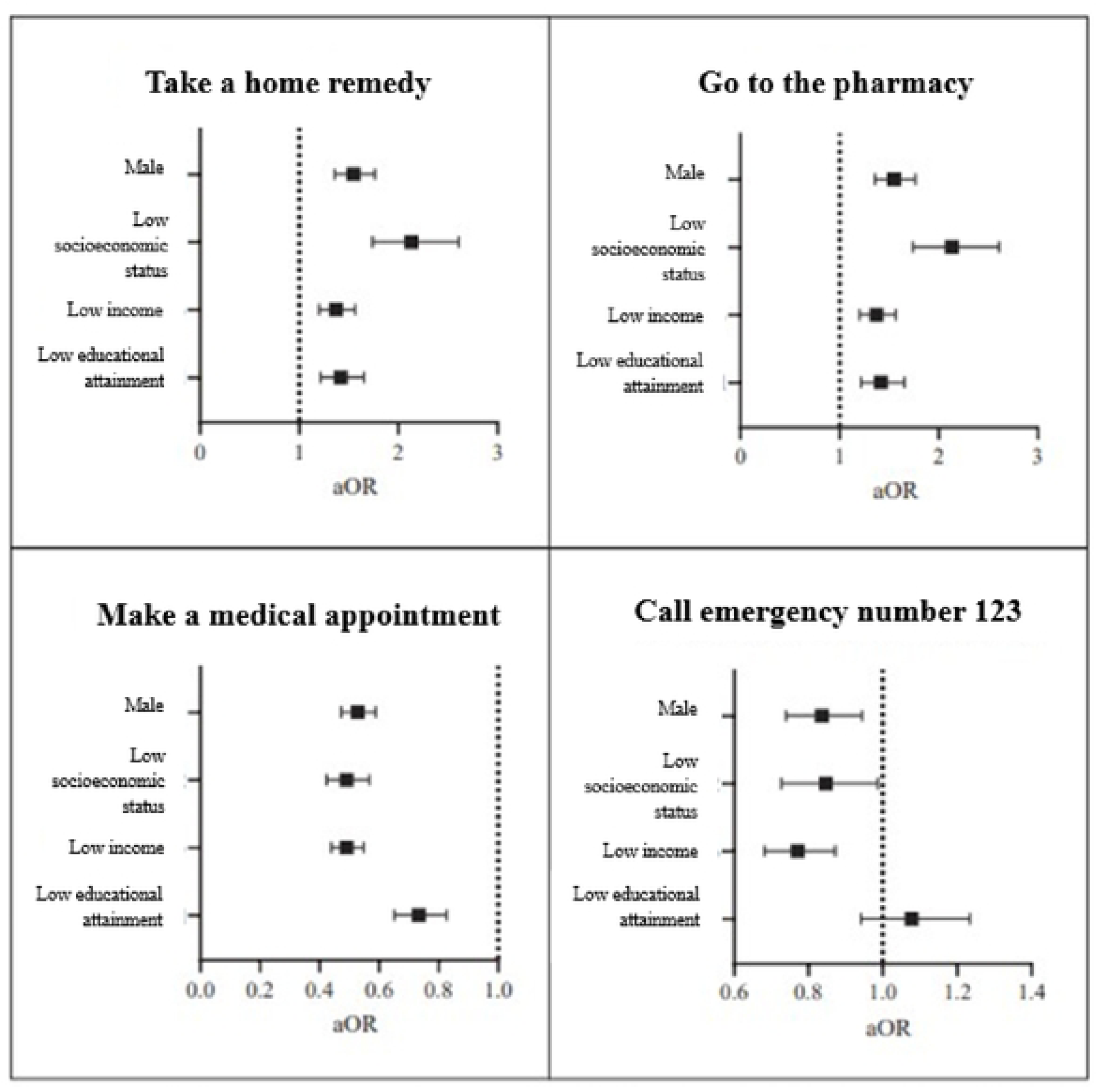
Predictive factors for stroke practices in the event of an acute Ischemic Stroke in Bogotá, Colombia. Summary of estimation of the association between stroke practices among participants and sociodemographic variables. Odds Ratio (aOR) was adjusted by variables of age and past medical history of systemic arterial hypertension.

## Discussion

This study provides information regarding the pool of knowledge, beliefs, and practices when confronting an IS from a large and representative population with vascular risk factors in an urban setting in Colombia. Despite being the second leading cause of death and one of the leading causes of disability in the world (1), the population’s perception of stroke risk and its impact on treatment access is a matter of controversy.

Data from the National Hospital Registry of Japan shows that there has been an increase in thrombolysis rates from 1.6% between the years 2000 to 2005, to 26.5% in the period between 2016 to 2020, with a comparable trend in endovascular therapy rates, increasing from 2% to 29.8% in the same period. (17) Similar trends have been reported in many countries worldwide. (18,19)

In Colombia, there is no data on this topic, neither national nor regional registries, but in most cases, they are single-center hospital registries, which do not reflect the national situation. The reported thrombolysis rates do not reach more than 17% for 2019, which suggests a much lower reality, even in regions with better access to reperfusion therapies. (20) Factors considered include the lack of recognition of stroke as an emergency requiring immediate attention, as well as socioeconomic factors and location in areas far from care centers. (21)

A systematic review on the topic in Africa showed an important delay from symptoms onset to hospital admission (31 hours), with poor knowledge of symptoms, not seeking medical attention, and shortage of medical transportation as the principal reasons for it. In India, Iran, and multiple countries in Africa, only between 8 and 30% of patients were admitted to a hospital in the first 3 hours from symptom onset. (22)

A population-based study conducted in Uganda in 2014, found that 71.8% of the participants did not recognize the brain as the affected organ in stroke and did not know any symptoms of the disease. Factors such as a high level of education or a diagnosis of type 2 DM were related as factors in favor of knowledge of the symptoms of the disease. (23)

Another population-based study conducted in New Zealand and published in 2020 found that factors such as a higher level of education, higher economic income, and personal experience of stroke were related to a higher level of stroke awareness. (24)

A study in a similar setting to Colombia, although population-based, is the EstEPA Project, carried out in Argentina by Dr. Ameriso et al. and published in 2021, who conducted a questionnaire to assess the level of awareness of stroke. A sample of 1986 responses was collected, among which 63% recognized stroke as a cerebrovascular accident. It was also found that the most common responses when the participants were worried about having an IS were going directly to the emergency room or calling the emergency services (52.3% and 39%, respectively). Factors such as age, sex, or race had no impact on the recognition of stroke to act accordingly. (25) In contrast, our study revealed that 73.4% of the population would call the emergency hotline. Furthermore, our findings highlighted a sex difference, as men were more prone to be unsure of the appropriate course of action.

The present study is the first conducted in the Colombian population and also the first in Latin America to address the knowledge and perceptions of the population at higher risk of having a stroke. Among the most important findings is that most of the people surveyed recognize stroke as a disease that can lead to disability (89.6%) and even death (91.1%). It is also interesting to highlight the recognition of the conditions they suffer from (HTN, DM, sedentary lifestyle, dietary factors, or smoking) as risk factors for suffering a stroke in a proportion that varied between 84.6% and 91.4% according to the factor, being quite similar to the proportion of people who recognize stroke as a condition that can leave sequelae or be fatal.

Furthermore, male sex, lower socio-economic status, and lower educational level were associated with lower knowledge of stroke symptoms. In the multivariate analysis, the same characteristics predicted a higher risk of choosing inadequate courses of action in the event of a stroke. There is, at least for this segment of the population, a gap between basic stroke knowledge and its application in real life.

Personal history of stroke and educational level have been shown previously to be predictors of better stroke knowledge in multiple studies, meanwhile, sex as a predictor of knowledge is a controversial topic as there have been contradicting results in different populations. (26–29) Looking for alternative solutions instead of going directly to the emergency department is common in lower-income countries. (30)

Ongoing efforts are underway globally to increase the number of patients receiving reperfusion therapies for stroke. This includes educating practicing physicians, increasing global access to EVT, and improving the capability of hospitals to treat stroke. (31) Nevertheless, it’s fundamental to include population awareness and education as an objective for these campaigns trying to increase treatment rates.

It has been shown previously that minorities and more vulnerable populations have lower chances of receiving reperfusion therapies, and one of the reasons could be a lack of stroke education and lower rates of prompt emergency department consultations. (32)

Another consideration to be taken into account by those responsible for the Colombian Health System, especially the Emergency Medical System, is the fact that almost 77% of patients consider calling the emergency care line to receive care in the event of a stroke. This reflects the urgent need to continue active work to educate about the performance of emergency services in the local environment, which to date has no published data, and thus breaks down the various barriers that may exist in practice and have not been addressed. In this regard, studies such as that of Ro et al. identified that the percentage of patients arriving less than 2 hours from the onset of symptoms was significantly higher if the patients arrived directly at the hospital brought by the emergency medical services, compared to those who arrived transported from another hospital that did not have the necessary resources to attend an IS. (33)

Educational campaigns have been shown to be efficacious in increasing stroke knowledge in different populations. (34,35) Whether this knowledge improves reperfusion therapies has yet to be widely studied, but some evidence suggests that it decreases the time from symptom onset to hospital arrival. (36)

The present study has several strengths. Firstly, it includes a large sample size collected from various regions across the city, making it the largest known study investigating health literacy about stroke. Secondly, it employs robust research methodologies, ensuring the reliability and validity of the findings. Thirdly, by focusing on non-health professional individuals, the study provides valuable insights into a segment of the population that is often underrepresented in health literacy research, which will inform and shape public health strategies and educational interventions aimed at improving stroke awareness and prevention. Additionally, the study represents a collaborative effort, involving multiple institutions and disciplines, providing a broad and comprehensive representation of Bogotá’s population in Colombia.

Regarding the limitations of this study, it is worth mentioning that the data were self-reported by participants, which may introduce bias or inaccuracies due to memory recall issues or social desirability bias. Additionally, the questionnaire used in this study had not been previously employed in similar research. Although it was validated by an expert committee, its novelty may still affect the consistency and reliability of the results. Moreover, the study included a multicultural and diverse population, which could result in variability in health literacy levels due to different cultural backgrounds, education levels, and health practices. This diversity can make it challenging to generalize the findings to a more homogeneous population.

## Conclusions

This research helps to better understand the reality of stroke in Colombia. It shows the necessity of high-quality educational measures regarding stroke symptoms, but most of all, to educate in early recognition and fast seek out of emergency medical services. Factors such as poor recognition of stroke as an emergency and socioeconomic disparities contribute to delays in seeking treatment. Addressing these barriers is crucial for improving stroke outcomes.

## Data Availability

All relevant data are within the manuscript and its Supporting Information files.

## Acknowledgments

Ana Barrera, Ana C. Martinez, Karen Calderón, Oscar Gil, Viviana Mercado, and Yeimi Trujillo actively participated in the study’s data collection.

## Declaration of conflicting interest

The authors have no potential conflicts of interest concerning this research.

## Funding statement

This study received funding from ATENEA, Agencia Distrital para la Educación Superior, la Ciencia y la Tecnología.

## Ethical approval and informed consent statements

Ethical approval for this study was obtained from the Institutional Institutional Ethics Committee (IEC) of the Government Social Entity (E.S.E.) North Health Services Subnetwork. Contact. Oficina Gestión del Conocimiento: Av. Suba #106-47 Sede Fray Bartolomé de las Casas - Subred Norte E.S.E. Bogotá, Colombia. Serial number: SNACEI-132. File 73. Written informed consent was obtained from all participants for their anonymized information to be published in this article.

## Notes

### Competing Interest Statement

The authors have declared no competing interest.

### Funding Statement

Yes

### Author Declarations

The Ethics Committee of the North Subnetwork of Health Services in Bogotá, Colombia, serves as an independent and multidisciplinary body responsible for safeguarding the rights, dignity, and welfare of individuals participating in health-related research. Operating under national regulations such as Resolution 8430 of 1993 and aligned with international ethical standards, the committee rigorously evaluates research protocols involving human subjects to ensure scientific validity, informed consent, and risk minimization. Its functions include reviewing methodological soundness, assessing ethical compliance, and granting formal approval prior to the initiation of studies conducted within affiliated institutions. By upholding transparency and ethical integrity, the committee plays a pivotal role in fostering responsible research practices and protecting vulnerable populations served by the subnetwork. Approval number: SNACEI-132. File 73.

## References

1. World Health Organization - Regional Office for the Eastern Mediterranean [Internet]. [cited 2024 July 25]. WHO EMRO | Stroke, Cerebrovascular accident | Health topics. Available from: http://www.emro.who.int/health-topics/stroke-cerebrovascular-accident/index.html

2. Global incidence, prevalence, years lived with disability (YLDs), disability-adjusted life-years (DALYs), and healthy life expectancy (HALE) for 371 diseases and injuries in 204 countries and territories and 811 subnational locations, 1990–2021 | Institute for Health Metrics and Evaluation [Internet]. [cited 2024 July 25]. Available from: https://www.healthdata.org/research-analysis/library/global-incidence-prevalence-years-lived-disability-ylds-disability

3. O’Donnell MJ, Chin SL, Rangarajan S, Xavier D, Liu L, Zhang H, et al. Global and regional effects of potentially modifiable risk factors associated with acute stroke in 32 countries (INTERSTROKE): a case-control study. Lancet Lond Engl. 2016 Aug 20;388(10046):761– 75.

4. GBD 2016 Neurology Collaborators. Global, regional, and national burden of neurological disorders, 1990-2016: a systematic analysis for the Global Burden of Disease Study 2016. Lancet Neurol. 2019 May;18(5):459–80.

5. National Institutes of Health (NIH) [Internet]. 2015 [cited 2024 July 25]. Health Literacy. Available from: https://www.nih.gov/institutes-nih/nih-office-director/office-communications-public-liaison/clear-communication/health-literacy

6. Nepal GM, Chand P, Poudel K, Acharya SR. Knowledge of Stroke among Hypertensive Patients in Dhulikhel. Kathmandu Univ Med J KUMJ. 2022;20(78):141–6.

7. Alhowaymel FM, Abdelmalik MA, Mohammed AM, Mohamaed MO, Alenezi A. Knowledge, Attitudes, and Practices of Hypertensive Patients Towards Stroke Prevention Among Rural Population in Saudi Arabia: A Cross-Sectional Study. SAGE Open Nurs. 2023;9:23779608221150717.

8. Cabezas RD. Conocimiento de síntomas y factores de riesgo de enfermedad cerebrovascular en convivientes de personas en riesgo. Acta Neurológica Colomb. 2015 Feb 25;31(1):12–9.

9. Cantillo-García V, Guzman LA, Arellana J. Socioeconomic strata as proxy variable for household income in transportation research. Evaluation for Bogotá, Medellín, Cali and Barranquilla. DYNA. 2019;86(211):258–67.

10. DANE - Socioeconomic stratification [Internet]. [cited 2024 Nov 14]. Available from: https://www.dane.gov.co/index.php/en/citizen-service/information-services/socioeconomic-stratification

11. Secretaría Distrital de Salud de Bogotá Reorganización del sector salud [Internet]. [cited 2024 Nov 14]. Available from: https://www.saludcapital.gov.co/Paginas2/Reorg_Sector_Salud.aspx

12. Salamanca VC. El Tiempo. 2023 [cited 2024 Nov 14]. 7 nuevos hospitales y 20 centros de salud le deja a Bogotá esta administración. Available from: https://www.eltiempo.com/mas-contenido/7-hospitales-y-20-centros-de-salud-le-deja-a-bogota-esta-administracion-763878

13. Sánchez MSZ, Sánchez CPZ, López PAC, Sanabria MS, Hernández SCH. Prevalencia de hipertensión arterial en Colombia: Acta Médica Colomb [Internet]. 2019 Sept 30 [cited 2024 Dec 12];44(4). Available from: https://www.actamedicacolombiana.com/ojs/index.php/actamed/article/view/1293

14. Page MJ, McKenzie JE, Bossuyt PM, Boutron I, Hoffmann TC, Mulrow CD, et al. The PRISMA 2020 statement: an updated guideline for reporting systematic reviews. BMJ. 2021 Mar 29;372:n71.

15. Guevara S. JD, Shields R. Spatializing Stratification: Bogotá. Ardeth Mag Power Proj. 2019 Mar 1;(4):223–36.

16. Chica-Olmo J, Sánchez A, Sepúlveda-Murillo FH. Assessing Colombia’s policy of socio-economic stratification: An intra-city study of self-reported quality of life. Cities. 2020 Feb 1;97:102560.

17. Koge J, Yoshimura S, Koga M, Nakai M, Wada S, Sasahara Y, et al. Nationwide Trends in Reperfusion Therapy and Outcomes of Acute Ischemic Stroke According to Severity: The Japan Stroke Data Bank. Stroke Vasc Interv Neurol. 2023 Nov;3(6):e000950.

18. Stein L, Tuhrim S, Fifi J, Mocco J, Dhamoon M. National trends in endovascular therapy for acute ischemic stroke: utilization and outcomes. J Neurointerventional Surg. 2020 Apr;12(4):356–62.

19. Fushida-Hardy N, Kim A, Leighs A, Thompson S, Tyson A, Barber PA, et al. Stroke reperfusion treatment trends in New Zealand: 2019 & 2020. N Z Med J. 2022 Mar 11;135(1551):68–80.

20. Moreno E, Rodríguez J, Bayona Ortíz H. Trombólisis endovenosa como tratamiento del ACV isquémico agudo en Colombia: una revisión sistemática de la literatura. Acta Neurológica Colomb. 2019;35(3):156–66.

21. Silva-Sieger F, Arenas Borda W, Zarruk Serrano JG, Restrepo J, Bernal Pacheco Ó, Ramirez Marroquín S, et al. Factores asociados al tiempo de consulta en pacientes con enfermedad cerebrovascular isquémica. Rev Neurol. 2007;44(5):259–63.

22. Urimubenshi G, Cadilhac DA, Kagwiza JN, Wu O, Langhorne P. Stroke care in Africa: A systematic review of the literature. Int J Stroke Off J Int Stroke Soc. 2018 Oct;13(8):797– 805.

23. Nakibuuka J, Sajatovic M, Katabira E, Ddumba E, Byakika-Tusiime J, Furlan AJ. Knowledge and Perception of Stroke: A Population-Based Survey in Uganda. ISRN Stroke. 2014;2014:10.1155/2014/309106.

24. Krishnamurthi RV, Barker-Collo S, Barber PA, Tippett LJ, Dalrymple-Alford JC, Tunnage B, et al. Community Knowledge and Awareness of Stroke in New Zealand. J Stroke Cerebrovasc Dis Off J Natl Stroke Assoc. 2020 Mar;29(3):104589.

25. Hawkes MA, Gomez-Schneider MM, Dossi DE, Melcon MO, Ameriso SF. Stroke Knowledge in the EstEPA Project, a Population-Based Study. J Stroke Cerebrovasc Dis Off J Natl Stroke Assoc. 2021 Feb;30(2):105471.

26. Alluqmani MM, Almshhen NR, Alotaibi RA, Aljardi OY, Zahid HM. Public Awareness of Ischemic Stroke in Medina city, Kingdom of Saudi Arabia. Neurosci Riyadh Saudi Arab. 2021 Apr;26(2):134–40.

27. Bhat V, Gs T, Kasthuri A. Stroke Awareness among Elderly Hypertensives in a Rural Area of Bangalore District, India. J Stroke Cerebrovasc Dis Off J Natl Stroke Assoc. 2021 Jan;30(1):105467.

28. Navia V, Mazzon E, Olavarría VV, Almeida J, Brunser AM, Lavados PM, et al. Stroke symptoms, risk factors awareness and personal decision making in Chile. A national survey. J Stroke Cerebrovasc Dis Off J Natl Stroke Assoc. 2022 Dec;31(12):106795.

29. Elshebiny A, Almuhanna M, AlRamadan M, Aldawood M, Aljomeah Z. Awareness of Stroke Risk Factors, Warning Signs, and Preventive Behaviour Among Diabetic Patients in Al-Ahsa, Saudi Arabia. Cureus. 15(2):e35337.

30. Owolabi LF, Nagoda M. Stroke in Developing Countries: Experience at Kano, Northwestern Nigeria. Sudan J Med Sci [Internet]. 2012 [cited 2024 Sept 10];7(1). Available from: https://www.ajol.info/index.php/sjms/article/view/78138

31. Saini V, Guada L, Yavagal DR. Global Epidemiology of Stroke and Access to Acute Ischemic Stroke Interventions. Neurology. 2021 Nov 16;97(20_Supplement_2):S6–16.

32. Man S, Solomon N, Mac Grory B, Alhanti B, Saver JL, Smith EE, et al. Trends in Stroke Thrombolysis Care Metrics and Outcomes by Race and Ethnicity, 2003-2021. JAMA Netw Open. 2024 Feb 7;7(2):e2352927.

33. Park HA, Ahn KO, Shin SD, Cha WC, Ro YS. The Effect of Emergency Medical Service Use and Inter-hospital Transfer on Prehospital Delay among Ischemic Stroke Patients: A Multicenter Observational Study. J Korean Med Sci. 2016 Jan;31(1):139–46.

34. Saini V, Guada L, Yavagal DR. Global Epidemiology of Stroke and Access to Acute Ischemic Stroke Interventions. Neurology. 2021 Nov 16;97(20_Supplement_2):S6–16.

35. Tan J, Ramazanu S, Liaw SY, Chua WL. Effectiveness of Public Education Campaigns for Stroke Symptom Recognition and Response in Non-Elderly Adults: A Systematic Review and Meta-Analysis. J Stroke Cerebrovasc Dis Off J Natl Stroke Assoc. 2022 Feb;31(2):106207.

36. Zhao J, Yuan J, Lu K, Rudd A, Liu R. Why we should raise stroke awareness in the younger population? CNS Neurosci Ther. 2023 Jan 11;29(3):757–9.

